# Explaining the outcomes of the ‘Clean India’ campaign: institutional behaviour and sanitation transformation in India

**DOI:** 10.1101/19004689

**Authors:** Val Curtis

## Abstract

**Introduction:** Whilst many less developed countries are struggling to provide universal access to safe sanitation, in the last five years India has almost reached its target of eliminating open defaecation. The object of this study was to understand how the Indian Government effected this sanitation transformation.

**Methods:** The study employed interviews with 17 actors in the Government’s ‘Clean India’ programme across the national capital and four states which were analysed using a theory of change grounded in Behaviour Centred Design.

**Results:** The *Swachh Bharat Mission (Gramin)* claims to have improved the coverage of toilets in rural India from 39% to over 95% of households between 2014 and mid 2019. From interviews with relevant actors we constructed a theory of change for the programme in which high-level political support and disruptive leadership changed environments in districts, which led to psychological changes in district officials, which, in turn, led to changed behaviour concerning sanitation programming. The Prime Minister’s setting of the ambitious goal to eliminate open defecation by the 150^th^ birthday of Mahatma Gandhi (October 2019) galvanised government bureaucracy, while early success in 100 flagship districts reduced the scepticism of government employees, a cadre of 500 young professionals placed in districts imparted new ideas and energy, social and mass media was used to engage and motivate the public and key players, and new norms of ethical behaviour were demonstrated by leaders. As a result, district officials engaged emotionally with the programme and felt pride at their achievements in ridding villages of open defecation.

**Conclusions:** Though many challenges remain, Governments seeking to achieve the Sustainable Development Goal of universal access to safe sanitation can emulate the success of India’s *Swachh Bharat* Mission.

**SUMMARY BOXES:** *What is already known?:*

- At least 47 countries are not on track to reach the Sustainable Development Goal of universal access to safe sanitation by 2030 and some 0.6 billion people are still defecating in the open.
- It is not clear how governments in low income countries can be galvanised to act to resolve this pressing public health problem.

*What are the new findings?:*

- The experience of the Clean India programme suggests that countries can almost eliminate open defecation.
- The success of the programme was due to factors including: the setting of ambitious targets; the use of modern communications strategies and monitoring technology; and the provision of visible reward and recognition for employees.

*What do the new findings imply?:*

- Disruptive leadership is needed to create working environments where sometimes jaded civil servants are given an opportunity to make a difference.
- Politicians who embrace the cause of sanitation may find that there are votes in toilets.

---

> *“In a country as powerful as India – dreaming of space conquest, building roads, development – the fact that the priority of the entire Indian administration is to end open defecation is extraordinary.”* (Interview #14)

## INTRODUCTION

In 2007 readers of the BMJ chose the introduction of clean water and sewage disposal—”the sanitary revolution”—as the most important medical milestone since 1840^1^. Yet safe sanitation, essential for the prevention of infectious disease, for child growth, for gender equity and for human dignity ^2 3 4 5 6 7 8 9 10^ has yet to reach a quarter of the world’s inhabitants^11^.

Most countries of South East Asia and Sub-Saharan Africa have struggled to improve their rates of toilet ownership. One country, however, has bucked this trend. Once responsible for 60% of the world’s open defecators^12^, over the last five years rural India has been transformed by a government-led programme named the Clean India Mission (*Swachh Bharat Mission (Gramin)* or SBM(G). In 2014, fewer than four in ten rural Indian households owned a toilet; by the middle of 2019, official figures put coverage at over 95%^13^. Whilst it is hard to be certain of the actual numbers of toilets in such a vast and diverse country, it is clear that a major national transformation has taken place. This is an unlikely success story, both because Indian bureaucracy is not famous for its ability to create rapid change, and because ending open defecation is a surprising target for a political campaign.

In this paper I aim to explain how this sanitation transformation happened. From interviews with national, state and district actors, I deduce a theory of change for the behaviour of government officials in districts and use this to draw lessons for other nations wishing to emulate the success of the Clean India campaign.

## METHODS

### Study sample

I interviewed seventeen individuals chosen to represent a range of types of SBM(G) actors operating at national, state, and district levels. Participants came from the capital and four states, with varied performance on toilet coverage. Of the interviewees, six were career civil servants, five were assigned to work in SBM(G), four were employed by partner organisations outside of government, and two were academics. Of the total, four worked in national government, four at state level and six at district level. The aim was to have approximately four individuals in each type of position (some interviewees represented several types) and to continue interviewing until saturation was reached^14^. Fifteen of the seventeen interviewees were Indian nationals and seven were female. Eleven interviews took place face-to-face and six over Skype; each lasted 60 to 80 minutes. Interviews took place between May and July 2018. All of the people approached agreed to take part, although one delegated the interview to a colleague. I also drew on my experience as an occasional adviser on behaviour change to the Indian Ministry of Drinking Water and Sanitation (MDWS).

### Study ethics

Interviewees were supplied with an information sheet about the study, were informed that their participation would be anonymous and were asked to sign a consent form. Identifying information was removed prior to transcription of interviews. Ethical approval was granted by LSHTM (no 15261) and permission to conduct the study was granted by the <u>MDWS</u>.

### Patient and Public Involvement

No patients or members of the public were involved in the design, analysis or reporting of this study.

### Data collection and analysis

The interviews followed a structured format that included a request to describe the SBM programme, questions about administrative structures, decision-making systems, programme financing, technical issues and specific challenges. I also asked participants about their own circumstances and their motives relating to their work in SBM(G). I followed COREQ guidelines for the design, analysis and reporting of qualitative research^15^.

Interviews were conducted in English and recorded. Following transcription, I employed the steps of framework analysis, namely: (i) familiarisation, (ii) identifying a thematic framework, (iii) indexing, (iv) charting, and (v) mapping and interpretation^16^. I employed the framework of Behaviour Centred Design (BCD)^17^ to index and chart the data using NVivo version 11^18^. Figure 1 shows the generic BCD Theory of Change framework, depicting how interventions perturb the physical, biological and social environment of actors, these changes affect their psychology (at an executive, motivated or habitual level), which leads to changes in their behaviour. These behavioural changes, when compounded, lead to changes in the state of the world, (for example, better toilet provision in India). These changes all take place in a particular geographical, political, economic, and cultural context.

**Figure 1.**
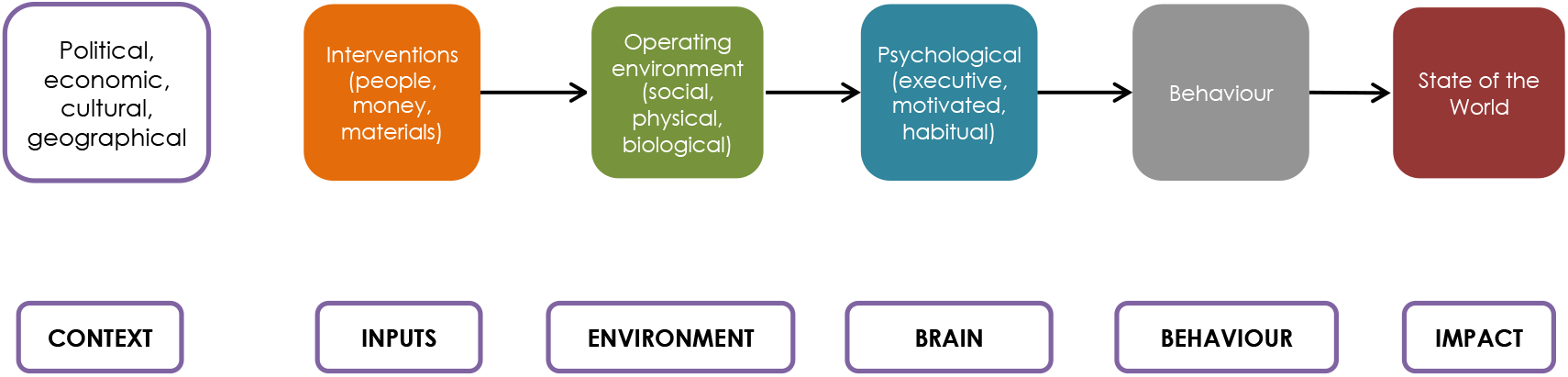
Generic Behaviour Centred Design Theory of Change

## RESULTS

Figure 2 shows the theory of change for SMB(G) constructed from the analysis of the interviews using the BCD framework. On the right are the observed changes to the state of the world (improved toilet coverage and use). Left of this, in grey, is the change in behaviour of district officials, in blue are the psychological changes in these actors, brought about by changes in their operating environments, shown in green. The orange boxes are the aspects of the intervention (i.e., the activities of SBM(G) as led by the Ministry of Drinking Water and Sanitation that caused this cascade of changes. Below I detail these changes, working from inputs to impact.

**Figure 2.**
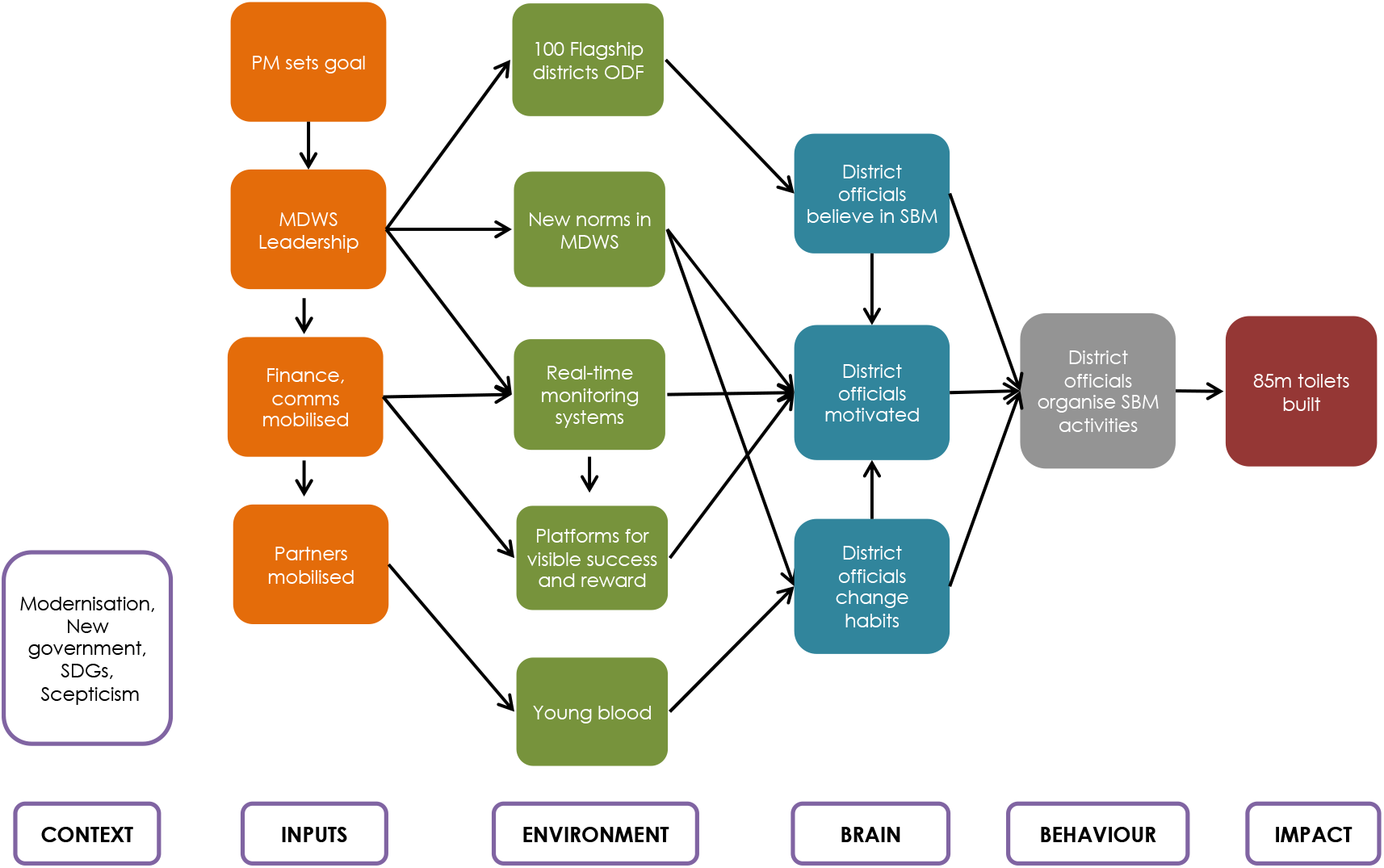
Theory of change for the *Swachh Bharat Mission (Gramin)*

### Interventions (inputs to SBM(G))

All interviewees ascribed the initial impetus for the *Swachh Bharat Mission* to the Prime Minister, Narendra Modi. In 2014, in his inaugural speech from the ramparts of the Red Fort in Delhi Modi announced his plan for India to become free of open defection in five years, by 2^nd^ October 2019, in honour of the 150^th^ anniversary of the birth of Mahatma Gandhi. The setting of this ‘Big Hairy Audacious Goal’^19^ made the need for radical changes in the government’s normal approach evident:

> *“[We are in] mission mode delivery arrangement, where you have a sunset clause that you have to achieve in a given time period… …So, it’s a very different working paradigm.”* (01)
>
> *“[In normal practice] the government might have said that we have 10 years to make India open defecation free. So we might have done squat for seven years and started working in the 8th year and then asked for a 5 year no-cost extension.”* (16)

Officials at all levels referred to the Prime Minister’s evident commitment to SBM:

> *“The major change is that the political leadership came from the Prime Minister’s office… …to reach all the communities, civil societies, religious leaders, famous people of India, actors, or people in sport. Involving everybody at the same time for a National Cause under the lead of the PM”*. (14)
>
> *“…the fact that this it is led by the Prime Minister – it is high on the agenda of every District Collector. It has led to government functionaries being on their toes all the time and also reaching out to the people in a way that has never happened before.”* (12)

And the PM kept up pressure over the life of the programme:

> *“So, it was not just a political announcement, but they backed it with serious attention over last four years, I think that’s quite dramatic. That’s one big game changer.”* (16)
>
> *“the PM referred to Swachh Bharat in 35 of his 50 addresses to the nation on All India Radio between October 2014 and Dec 2018”* (10)

Breaking with administrative norms to appoint staff on the basis of seniority, the Prime Minister sought out a technocrat to lead SBM(G) within the Ministry of Drinking Water and Sanitation. The chosen Secretary was a sanitation specialist and was also unusual in having worked both within the Indian Administrative Service, and outside of it in international bodies. This gave him the ability not just to navigate the government hierarchy, but also to see where it might be disrupted.

> *“In terms of technical leadership, this time the people put in place to lead this campaign are people with global experience that have worked abroad. They are aware of what is happening in the world in terms of sanitation programming, successes and failures, so there is great openness to learning in this program.”* (14)

The ministry team led a consultation exercise with States to gather lessons from a succession of previous sanitation campaigns, and to work out what it would cost to achieve the goal of an ODF India. The total bill was estimated at some $20bn over 5 years, or about $4.00 per head of the population per year^20^. In addition, the team negotiated a loan of $1.5bn from the World Bank^21^. The ministry also asked the World Bank, UNICEF, WaterAid and other support agencies to augment their technical assistance to states, and asked philanthropic foundations (Tata Trusts and the Bill and Melinda Gates Foundation) to fund the recruitment of consultants to a project management cell:

> *“The consultants have given a new dimension to the working of the Ministry. Previously we were working like a Government setup, but with the coming of this young blood… …they have given momentum to the program*. (02)

Unusually for India, the management cell was integrated within the Ministry rather than sitting outside of it. The SBM(G) team thus had access to government communications channels as well as to the cabinet and Prime Minister:

> *“What Modi has done is picked up these bureaucrats and said you have direct access to me*…*”* (09)

A further input that distinguished SBM(G) from standard Indian government programmes was a new strategy for communications. The team recognised early on that it was important to constantly communicate; downwards through the government hierarchy to programme actors, outwards to the population as a whole and to partners in the private and development sectors, and also, critically, back upwards to SBM’s political masters. Key influencers had to be kept on board:

> *“We started delivering and then we started communicating that to the people who matter - like media, like the political masters, they need to feel that the thing is moving. Then there are academics, the international community, the sort of people who are influencers, who can spread the word. It’s like chicken and egg: when you start doing well then nothing succeeds like success, but it needs to be communicated*…*”* (11)
>
> *“So point is if you want political leadership and you get it, then you have to nurture it…”* (14)

Mass media played a <u>major</u> role in keeping all of these audiences supportive and engaged:

> *“…all of our communication has become much more world class. Indian Government ads are [normally] easily identifiable from their look and feel -- they are dull, boring and preachy and nobody wants to watch them. But in this ministry the ads are very exciting and entertaining*…*”* (10)
>
> *“Honestly I don’t know if promoting stuff on social media is leading to behaviour change in rural India, but it does make urban India more aware about what happening in rural areas, for that is also important”*. (12)

### Changes to the environment in districts

The Theory of Change depicts how these inputs (disruptive leadership, mobilisation of human and financial resources, new modes of communication) changed the operating environment for staff working in India’s more than 600 districts. The new leadership team in the Ministry recognised that the first barrier to overcome was widespread staff scepticism, engendered by the experience of having seen many previous campaigns falter^22^. They needed to make district officials believe that change was possible. One hundred districts were therefore targeted for an ‘early-win’ campaign. Their heads, the District Collectors, were invited to national workshops, and were offered resources and, unusually, direct access to the leadership, so as to be able to resolve any problems they encountered. By the end of 2016, most of these flagship districts had declared that they were Open Defaecation Free (ODF). This model was then duplicated across the country.

> “*That is one of the great achievements [of SBM], they really have made sure that there is great support for any collector who wants to achieve this.”* (17)

The new leaders of SBM disrupted social norms in districts by modelling behaviour that was results-oriented and ethical. One interviewee recounted:

> *“If the Secretary is going to the field [normally], the collector’s job is to be the protocol officer, to receive him at the airport, ensure his dietary requirements, see his arrangements are proper. Your job is to carry his briefcase. But this Secretary focuses on the task in hand, he asks the collector important questions, rather than where to buy this handloom saree for his wife… This changes the professional ethics. Then the administrators know that this guy will ask for results rather than where to go shopping.”* (09)

Several interviewees told of the surprise that they felt when they saw a photograph of a top ministry official climbing into a toilet pit to empty it in the national newspapers^23^. One top state official dug toilet pits herself. Another related her norm-violating initiation to the SBM programme by a trainer:

> *“‘Tatti’[shit] is the most disgusting and vile word in Hindi. And he was just like: ‘tatti tatti’… …and I could not stop giggling. It is not even a word you may use, you may use this word at home, but not if there is a guest or someone.”* (10)

Many interviewees felt that such activities had caused a change of norms in society, and by extension, in districts:

> *“So this is quite unique… …five or six years back you could not even talk about open defecation. And now you see that the whole of India is expert on the issue of open defecation.”* (14)

A further change to the operating environment of districts was the use of technology to monitor and encourage progress. Collectors were set targets by the leadership in Delhi and then, unusually, were then held to account. They were expected to report on progress in face-to-face meetings, in regular multi-participant video conferences and online.

> *“Video and satellite conferences were my two important communication tools to connect with people and officials from across all the districts. I used video conferences to get figures on how many toilets had been built and to handout targets*.” (15)

District Collectors were trained on and encouraged to sign up for social media, and to use email:

> *“[It is] rare for a bureaucrat to reply within an hour to an email. That means he has got email on his phone*. Babus *[bosses] still don’t do it to this day.”* (10)

In addition, the MDWS created a live ‘dashboard’; a website displaying progress on sanitation coverage showing the number of toilets built and the percentage of households covered ^24^. In many states this system worked automatically via a mobile phone app that was updated by village authorities in real time, as toilets were built and funds disbursed to householders’ electronic bank accounts.

These improvements in communications technologies both provided an environment where districts knew they could be held publicly to account, but also allowed districts an opportunity to show off their success. Rather than being forgotten in remote corners of the country, those collectors and their staff who did well were held up as examples on social media, invited to receive awards at multiple ceremonies involving local, state or national dignitaries and some were given awards by the prime minister himself.

> *“The secretary… …travels a lot to see what’s happening on the ground and he tweets almost every day about the good things that are happening. So, this provides a lot of motivation to the system.”* (08)
>
> *“We had this workshop to felicitate the spouses which really touched all of them. Many of these wives… …said that nobody has ever acknowledged the work that we do.”* (15)

A further means by which ‘business-as-usual’ in districts was disrupted was the introduction of ‘fresh young blood’. Aside from deploying technical consultants, development partners were asked to recruit a cadre of new, young and enthusiastic fellows. Five hundred *preraks* were hired and assigned to support districts, where they took on a variety of roles.

> *“This is how the program works: they take fellows from districts which have become ODF and put them in districts or states wherever it’s required according the skills and the needs*…*”* (08)
>
> *“The* preraks *are an astonishing innovation… They have been terrific…”* (17)

*Preraks* described how they offered flexible support to districts, filling in where needed:

> *“I had a collector who would call me pretty much on daily basis. If I didn’t meet him, he would text me and ask about the update. Eventually, I would run a lot of workshops and he even sent me to different locations because [he said] ‘I can’t understand what the problem is there, why don’t you go and check out what the problem is and just address it*…*’”* (07)

*Preraks* supported each other and shared solutions to problems, often using social media:

> “*But in terms of everyday things that we needed to carry out, we were calling each other on WhatsApp or email… ‘I heard that you guys have a lot of dysfunctional toilets, how are you working on this?’ …that was on a WhatsApp group*.” (08)

### Psychological changes

In figure 2 innovations that changed the social and technological environments in districts led to psychological changes (belief, motivation and habitual response) in the relevant officials. First, staff had to *believe* that change was possible. Officials repeatedly described how they had joined a civil service out of a desire to ‘serve’, but had been frustrated by bureaucracy and stasis:

> *“It’s just pushing up against the wall which is not going to move ever. So, why am wasting the good years of my youth and all my energy and ideas on this?”* (10)

But as they saw progress gathering in SBM, staff begun to believe that the sanitation situation could be changed:

> *“Because it gives you so much conviction, you know, what the world thought is impossible is being done in my time.”* (01)

As the results of their work became more visible, district officials related their pride at their achievements:

> *“So now when I travel with my family they say: ‘see what is happening to your Swachh Bharat…’…I am ‘Swachh Bharat Champ’ they call me.”* (03)

Staff described their motivation for working in SBM using emotive terms:

> *“There is a lot of passion for the first time in my sanitation business for many years. This is the first time that I have seen such a powerful country being mobilized in such a genuine and realistic way.”* (14)
>
> *“…it is glamorous”* (07)
>
> *“…disgust with the kind of situation people have lived in in our villages for such a long time…* (06)
>
> *“I think this whole country has been amazingly galvanized [by this] compelling creation, amazing euphoria…, …the sanitation program is a very adventurous, courageous and romantic venture.”* (01)

However, the motive that was most often mentioned by staff working in districts was the opportunity that working in the SBM had given to them to ‘make a difference’:

> *“It gives you satisfaction when you see that we are heading a program that is being implemented and that is being successful and people are owning it.”* (03)
>
> *“Because here I think I can give my best output and can contribute in assisting my bosses so that they can do it well too”* (02).

The new technological environment also changed the habitual responses of officials, for example, by making it easy for them to monitor progress closely:

> *“So now when you are in a village you log into the app [and] all the Swachh Bharat mission toilets are mapped there with the beneficiary name, with the photographs, everything is there.”* (06)

Further, the new norms of behaviour set by national officials led to new working habits such as abandoning air-conditioned offices and spending more time in the field:

> *“And then the districts which are very behind I visited all of them… I most often visit those districts which are in need.”* (03)

### Changes in behaviour in districts

The cascade of changes in the operating environment and in the psychology of actors in SBM led to new behaviour in districts – the level where staff have the mandate, operational staff and budgetary autonomy to organise development activities. Interviewees reported that the most important determinant of progress in latrine construction was the level of engagement of the District Collector (DC). A DC who had taken on the challenge of SBM would typically move into ‘campaign mode’, placing building toilets at the head of her/his priority list, sometimes neglecting other activities to do so. Components of the work included reviewing the sanitation status of blocks and villages, setting targets for toilet construction, organising payments to self-help groups and contractors, training masons and huge numbers of social mobilisers (*Swachhagrahis*), organising mobilisation events, monitoring results and verifying the open defecation-free (ODF) status of villages.

DCs used a range of strategies to galvanise these activities; setting targets for blocks (administrative units) and villages to become ODF, and for disbursement of funds, having weekly problem-solving meetings with block staff and sometimes following up daily on progress using social media.

> “*The commitment of some of these collectors is quite extraordinary. Some use WhatsApp to call people at 4 in the morning to make sure they are up for the nigrani samiti [morning visit to open defecation grounds]. Quite amazing!”* (17)
>
> *“So they [block coordinators] feel happy and motivated as there is regular follow up. And as I call them almost every day, so they also hold meetings almost every two to three days.”* (03)

Perhaps the biggest change in the behaviour of district level staff was the move from ‘business-as-usual’ to a personal engagement with the cause:

> “*If you are not out in the field, you are not present and people don’t see you, then people will not get excited.”* (11)
>
> *“Everybody was more excited about the mission, because the collector himself was sitting with us*.” (08)
>
> *“So you try to inspire people, motivate people, appeal to them, persuade them to do it fast, quickly and so on, in the mode of pulse polio [national eradication campaign], where in matter of a week or 10 days you can simultaneously start the work of construction and of triggering people.”* (15)

### Change in the State of the World (impact)

What, finally, was the overall impact of these changes in the administrative system under the *Swachh Bharat Mission (Gramin)?* The Government of India has made repeated efforts to improve the sanitation conditions of its rural population. Fig 3 shows the results. After the national census of 1981 put national sanitation coverage at only 1%, the Central Rural Sanitation Program was launched in 1986. This increased coverage to 9% and evolved into the Total Sanitation Campaign which began in 1991. In the following 20 years, coverage improved by some 1% a year, reaching 31% by 2011^25.^ Despite a relaunch of the campaign, as the *Nirmal Bharat Abhiyan*, India failed to meet its Millennium Development Goal to halve the rate of those with no access to sanitation by 2015^26^.

**Figure 3:**
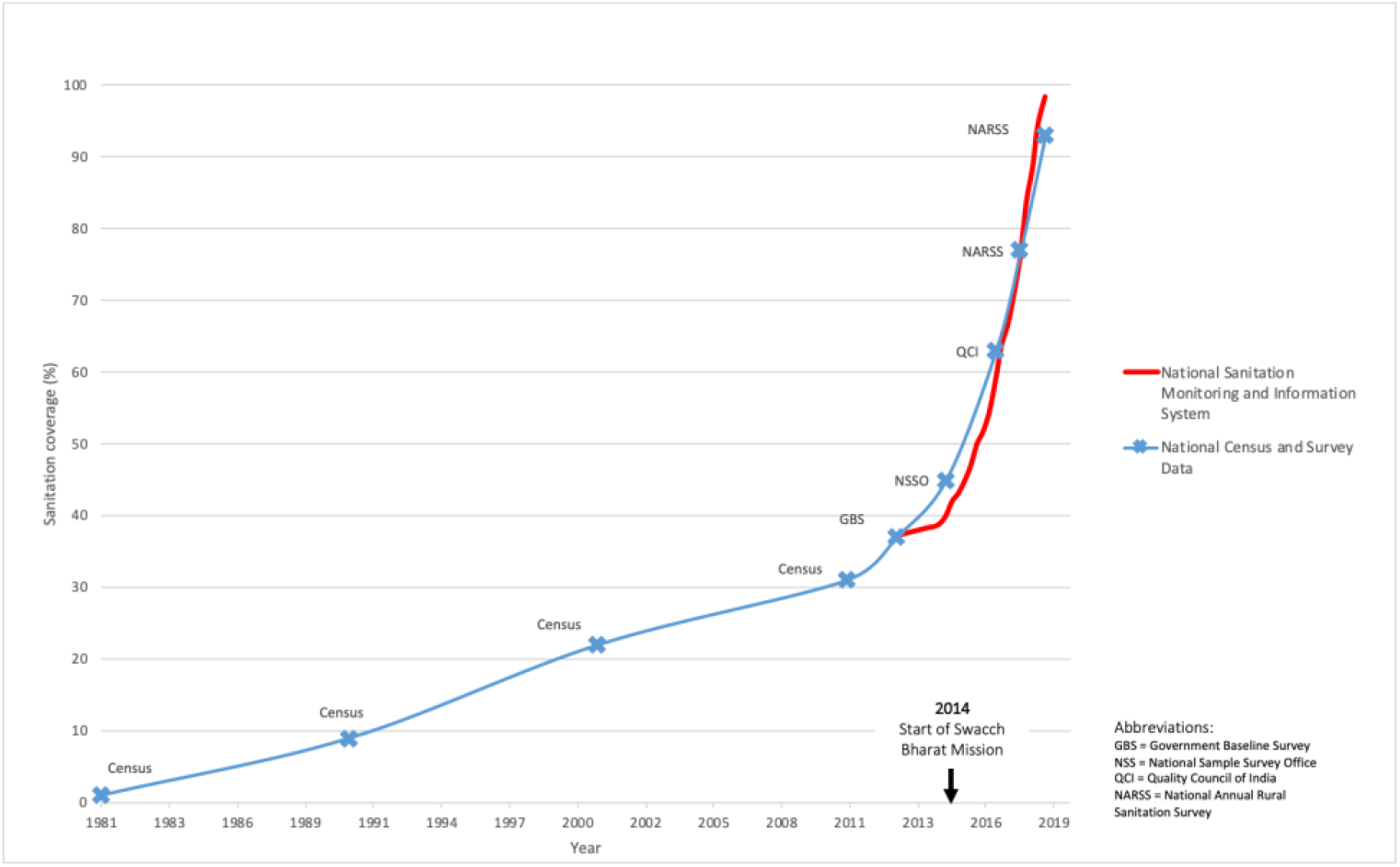
Household toilet coverage in India 1981-2018 according to surveys and the national monitoring system

By contrast, four years after the 2014 launch of *Swachh Bharat* official figures suggested that over 80 million toilets had been built, 500 districts and almost half a million villages had been declared to be free of open defaecation^27^. Figure 3 suggests that the rate of toilet building in rural areas was around ten times that of previous campaigns. In tandem with accelerating increases in sanitation coverage, government figures suggest that open defecation has also decreased, dropping by 3% per year between 2000 and 2014 and by 12% a year from 2015-2019^11^.

A number of unpublished surveys suggest that the government monitoring system overestimates the true levels of toilet coverage^28 29^. Interviewees gave reasons for discrepancies between individual surveys and government figures that included inaccuracies in the original government baseline against which progress is reported, the omission from the SBM programme of households with old toilets which had fallen out of use, and systemic incentives to overreport results. Nevertheless, interviewees described exceptional progress in improving toilet coverage over the past four years, transforming India from a country where the majority defecated in the open, to one where the majority of people have, and use, toilets.

> “*I don’t think there is any historic parallel of any country anywhere in the world, shifting numbers on such a scale. That will change the global indicators. I think that the story is not whether it is 85%, right, or 90%, right, the story is the fact that there is a tremendous shift*” (16)

Whilst interviewees were positive about the gains made by the SBM, there were also worries about the sustainability of these gains over the long term:

> *“we brought India from A to B which is an outstanding achievement but there is recognition that behaviour change is complex, there is unfinished business and there is necessity to invest beyond the campaign so that we have a real impact by the end of the SDG era”* (14)

As the final deadline for SBM looms, political attention to the unfinished last mile on sanitation will inevitably shift to other pressing social issues such as rural water supply. In the face of this the MDWS are trying to embed sanitation in national policy and plan to continue to make funds available to complete and sustain the gains of SBM.

> *“On sustainability, of course the ministry is also aware; they have started focusing on something called ODF Plus, which looks at the sustainability issue.”* (13)

### Context

Drawing transferable lessons from a programme’s Theory of Change requires understanding its particular context^30^. The SBM(G) programme took advantage of opportunities presented by the prevailing national political situation, of improvements in living conditions in rural India and of the proliferation of communications technology. It also drew on the experience of previous sanitation programmes, and on the global push to meet the Sustainable Development Goals. Contextual factors that hindered progress, on the other hand, included a general scepticism of government sponsored programmes, political division, where some states were not supporters of the current Government, and the many other competing demands for social investment in India.

Interviewees pointed out how the political context was key in the launch of SBM, where a political swing towards nationalist and pro-Hindu party provided an opportunity to capitalise on the symbolism of purity, cleanliness of the mother country and the example of Gandhi. This was expected to translate into electoral support:

> *“Sanitation is something each family needs every day. And through his program, Modi is able to reach out to the almost to all the households in the country. And a toilet becomes synonymous with Modi. So, Modi is equal to Swachh Bharat and Swachh Bharat is equal to a toilet. So, this has given him immense visibility, immense publicity and immense connection with crores of households in India.”* (01)

Indeed, observers have suggested that the Prime Minister’s evident commitment to rural sanitation was a factor in the success of the ruling party at the 2019 general elections^31 32^.

> *“You see, good sanitation is good politics.”* (01)

The new SBM goals were set in the context of a nation in the <u>midst</u> of modernisation, where millions have emerged from poverty, and most people have improved their housing and gained access to electricity, water, mass communications and education^33^. India was therefore ready for modern toilets, and had the communications networks necessary to rapidly spread this idea.

The SBM(G) programme did not emerge *de novo* but was set up in the context of a long line of previous national sanitation campaigns. These had met with limited success, but had provided experience concerning what worked and what did not.

> *“The Swachh Bharat mission is preceded by several other missions; the Total Sanitation Mission, Nirmal Bharat Abhiyan and other campaigns. When the Swachh Bharat mission started we were very much aware of this. We can’t say that those programs were a total failure, these contributed to building momentum and creating awareness, and some of these toilets are still in use. …definitely they left imprints.”* (05)

Whilst the national context was favourable to SBM, so was the international context. International agreement to meet the SDGs reminded Indian leaders that, though their country had made progress on many issues, sanitation conditions still lagged far behind that of countries with comparable levels of development.

## DISCUSSION

### Principal findings

Whilst it will still take some time to completely realise the ambition of an open defecation free India, the country has made large strides in this direction over the last five years. In rural areas of other low income countries there are still some 1.5 billion people who lack access to basic sanitation services and 0.6 billion who defecate in the open^34^. This study offers insights and lessons for the governments of countries that are looking to improve the sanitation conditions of their populations:

1. *There are votes in toilets*. The Indian experience suggests that political engagement at the head of government can galvanise an administration to a goal as unlikely as ending open defecation, and that this may be rewarded politically
2. *Disrupt institutional norms*. From the outset, the leaders of SBM(G) set out to disrupt the bureaucratic norms of government. The leadership modelled ethical norms of commitment to progress, new enthusiastic young staff in districts helped to create norms of adaptive learning, enthusiasm and personal engagement and the norm of polite silence regarding matters faecal was repeatedly and deliberately violated
3. *‘Believe the impossible’*. A key driver of the success programme was the ‘Big Hairy Audacious Goal’ to make India ‘Open Defecation Free’ by October 2^nd^ 2019. Standard development programmes tend to be conservative; setting less risky but more plausible goals (for example, the Tanzanian Government has set a target of 85% coverage with improved toilets by 2030). However, such targets neither provide a compelling vision nor a sense of urgency. SBM shows other countries that ambitious plans for sanitation transformation can galvanise behaviour change in institutions.
4. *Set targets and monitor them*. Many development programmes set targets, but few follow them up as relentlessly as did the *Swachh Bharat* team. Making use of electronic data gathering and real-time, public dashboards, the ministry team reviewed progress district by district, every month, sought to resolve any bottlenecks that emerged and held district collectors to account for their results.
5. *Reward and recognition*: Once districts began to meet their targets, their progress was deliberately and publicly celebrated. Officials from successful districts were lauded and presented with awards at national, state and local events, and they, in turn, gave awards to leaders and mobilisers in successful villages. Successful officials were praised in tweets and Instagram posts. Partly as a result, officials were full of pride at their achievements and were motivated to make efforts that went well beyond their normal pattern of work.
6. *Constant 360*^*0*^ *communications*. Unusually for a government programme, the SBM(G) embraced modern communication strategies and media technologies. The centre kept relevant stakeholders, such as MPs, journalists, academics, development partners and private sector players informed and engaged, using social media, mass media, high profile events, conferences, teleconferences, and personal visits. District officials learnt to use communications technologies such as email and social media to follow progress, to short-circuit sometimes cumbersome official channels and to encourage one another.
7. *Focus on behaviour and sustainability*. Learning from previous programmes in India an elsewhere, the SBM made major efforts to support behaviour change through mass training programmes and local innovation.
8. *A passion for sanitation*. A surprising finding from this research was the degree of emotion shown by the respondents when they described their engagement with the SBM programme. Motives driving their participation included justice, creativity, nurture, status and disgust^35^. From top to bottom of the hierarchy, almost every interviewee described the programme in emotive terms; whether as adventurous, glamorous, exciting, fulfilling, pride-generating, satisfying, or humbling. According to one interviewee:

> *“I will be the living testimony to the fact that India will become ODF and I will be making huge difference in lives of millions of people, because I am associated. The difference that we will make in rural India, the profound impact it will have on the lives of the poor people is very, very satisfying, inspiring, humbling.”(01)*

Whilst much has been written about many aspects of sanitation programming, such as the choice of technology^36^, motives for the adoption of toilets^37 38^, provision of subsidy^39 40^, community participation^41 42^, environmental factors^43^ and health outcomes of sanitation programmes ^44 45 46^, only a few published studies have tried to unpick the successes and failures of Government-led programmes.

A 2012 study of the of the previous Total Sanitation Campaign in India^22^ blamed its poor results on a lack of political priority and leadership, a lack of confidence in the possibility of success, the misuse of subsidies, poor monitoring systems, and a top-down supply-led approach. The results of the current study suggests that SBM(G)’s leaders had learnt how to resolve at least some of these problems; capitalising on political support, broadcasting the successes of the programme, using electronic banking to pay subsidies direct to households, employing technological platforms for monitoring and by emphasising behaviour change. The use of electronic technology for progress monitoring is not new to India but has been employed successfully in sanitation programmes in Indonesia ^47^ and Zambia ^48^. Whilst lack of political will has long been lamented in the field of sanitation (for example in India ^43^, Nepal ^49^ Tanzania ^50^ and Ghana ^51^) the SBM example suggests a reason why politicians should engage – because there are votes in toilets.

The factors that were responsible for the success of SBM to some extent echo those that have been identified as important for success in large scale health programmes more generally. One survey of 20 proven programmes concluded that political will, technological innovation, expert consensus around the approach, management that effectively uses information, and the deployment of sufficient financial resources were critical ^52^. It also pointed to the importance of Government ownership of development programmes if success is to be sustained.

Whilst the setting by the Government of ambitious, ‘not-quite achievable’ big hairy audacious goals was undoubtedly one of the factors in the success of the SBM, the pursuit of a success narrative can have what Rajkotia describes as an ugly side: “the high stakes, the ambition and the expectation can instil a fear of failure, stifle risk-taking and innovation, and lead to the fabrication of achievement.”^53^ In the case of SBM a high level of awareness of previous failures actually led to innovation, for example in an attempt to focus on behaviour change as well as on construction targets. The success narrative has also undoubtedly led to inflated claims about SBM’s results. But, as Rajkotia also points out, aspirational targets are important because they can rally governments and civil society to focus their energies on social development. He suggests that it might thus be unfair to chastise them for failure to completely achieve what was aspired to. For the future, countries should recognise that there is a virtuous circle by which aspirational targets can drive success, both through success itself, and by the learning that accrues from acknowledging, embracing and understanding failure to achieve them, partially or completely^54^.

Even when toilets have been built, and counted, individuals may still not use them. A study in Orissa suggested that whilst the Sanitation Campaigns of 1999-2012 increased latrine coverage substantially, over a third of people with toilets were still not using them ^55^. Though getting good data on usage is difficult, there are indications that the longer people have toilets the more likely they are to use them^56^. Evidence suggests that of all the structural, cultural, psychological and material factors determining toilet acquisition and usage the most important seems to be the perceived social norm of not defecating in the open^38^. Given the much wider presence of toilets and the norm-changing national conversation about them that has been engendered by SBM, it may be that toilet use has reached a tipping point, a point beyond which toilet use will only increase in India. This open question requires vigilance from government and close research attention in the coming years.

A novel aspect of this study was the attention paid to the settings and motives of civil servants. Whilst this study documented many positive emotions associated with participation in SBM(G), negative emotions have also been documented. Coffey and colleagues suggest that a consequence of the pressure to achieve complete toilet coverage in SBM(G) has been that some officials fear losing their jobs if they do not perform (Coffey, unpublished). In Bangladesh, however, Hanchett found that a factor in the success of the national sanitation programme was the enthusiasm and pride of union council chairmen, and experience sharing among them^57^.

### Strengths and weaknesses of the study

Courtesy bias may be partially responsible for the fact that interviewees were almost unanimously positive about their experience of SBM. However the enthusiasm of the SBM participants was clearly genuine. Given the huge size and heterogeneity of India, a larger study with a broader range of interviewees from a broader range of states might have provided further lessons, however, data saturation was achieved here with few new insights emerging after about 12 interviews^14^.

This study was unusual in that it focused on the behaviour of district officials within government institutions, rather than on the effect of sanitation programming on villages and households. A more complete analysis might have analysed the factors leading to behaviour change at every level in the hierarchy, from leaders, through Ministry, State, district, block and village, through to householders. A future analysis of the factors leading to behaviour change at the level of households in this programme could be expected to provide further rich findings.

The study also focused purely on sanitation programming aimed at households in rural India. How to achieve universal access to sanitation in urban settings in low income countries remains a large and largely unsolved problem.

Looking at the context in which SBM achieved what it did, it may be asked how far is India a special case, and how transferable are the results to other countries? India is one among many countries in which modernisation is proceeding rapidly, Tusting et al show that housing is improving across sub-Saharan Africa^58^, and our research in Tanzania suggests that toilet promotion can be linked to this process of modernisation^59^. India is also one of many countries in the Global South with political leaders with transformative ambitions, as was demonstrated when 53 government delegations attended a summit on SBM held in Delhi in October of 2018. India is different from many countries with unfinished sanitation agendas in its low level of reliance on external budgetary assistance. Yet the, seemingly huge, amount it made available for toilet building only amounted to some $4 per head per year, a sum that could be recouped in financial savings associated with lower health care and other costs^60^. Case studies suggest that poverty is rarely a barrier to the achievement of public health goals^52^. Hence it seems likely that other countries with the will to do so can emulate India’s sanitation success.

Employing the Theory of Change perspective of Behaviour Centred Design, which was here applied for the first time to behaviour change in institutions, provided a new, relatively straightforward and insightful tool for understanding the behaviour of actors in government. A novel aspect of the approach was the ability it gave to investigate the role of psychological factors such as motives, as well as the social and physical factors in the operational environment of the key actors. It allowed the creation of a structured and plausible theory of change from which it was possible to derive lessons that should be of use to other institutions desiring to make a step change in the quality and impact of public services.

### Issues and future research

Whilst SBM has measured its success against a baseline set in 2013, interviewees pointed out that many families had been left out of SBM. The Government will need to tackle the tough ‘last mile’ issue of remote and left-out rural households, as well as to find better sanitation solutions for the urban poor, before it can fully claim to have met its objective to declare the country free of open defecation. Questions remain concerning the sustainability of the achievements of SBM. Much could be learnt by studying the different experience and differing levels of success across India’s widely varied 600+districts. Future studies, including the national census of 2021, will reveal the long term trajectory of the gains claimed by SBM.

Finally, it is increasingly understood that individuals working in institutions are motivated to act by a range of factors beyond immediate financial advantage^61 62^. As one interviewee said:

> *“All of us who are members of civil services, we joined the service with the vision for the betterment of society.”* (05)

A closer investigation of the pro-social motives of civil servants in national action programmes such as the successful Swachh Bharat programme in India might reveal important lessons about how best to stimulate and reward such activity. This could have consequences beyond the urgent struggle to get safe sanitation to everyone on the planet.

## Data Availability

Transcripts are not available because ethics requires anonymity

## Abbreviations

COREQ: COnsolidated criteria for REporting on Qualitative research
DC: District Collector
ODF: Open Defaecation Free
MDWS: Ministry of Drinking Water and Sanitation
SBM: Swachh Bharat Mission
SBM(G): Swachh Bharat Mission (Gramin)

## Sources of funding

London School of Hygiene & Tropical Medicine

## Competing interests

The author declares no competing interests

## Acknowledgements

I wish to thank Robert Aunger, Sarah Bick, Kavita Chauhan, Ian Ross, Astrid Thorseth and the interviewees.

## Notes

### Competing Interest Statement

The authors have declared no competing interest.

### Clinical Trial

n/a

### Funding Statement

No external funding was required for this work. The author's salary was supported by LSHTM

### Author Declarations

All relevant ethical guidelines have been followed and any necessary IRB and/or ethics committee approvals have been obtained.

Any clinical trials involved have been registered with an ICMJE-approved registry such as ClinicalTrials.gov and the trial ID is included in the manuscript.

